# Analysis of potential barriers for non-PrEP users among MSM in Germany

**DOI:** 10.1101/2025.08.29.25334713

**Authors:** Helena Waldorf, Ulrich Marcus, Sara Iannuzzi, Stefan Albrecht, Jens Hoebel, Barbara Gunsenheimer-Bartmeyer, Viviane Bremer, Max von Kleist, Uwe Koppe

**Affiliations:** Project group 5 “Systems Medicine of Infectious Disease”, Robert Koch Institute, Berlin, Germany; Department of Mathematics and Computer Science, Freie Universität Berlin, Berlin, Germany; Department of Infectious Disease Epidemiology, Robert Koch Institute, Berlin, Germany; Department of Epidemiology and Health Monitoring, Robert Koch Institute, Berlin, Germany

**Keywords:** HIV prevention, barriers, PrEP, TDF/FTC, MSM, uptake

## Abstract

**Background:** Daily oral pre-exposure prophylaxis (PrEP) provides effective protection against HIV. Since September 2019, the costs of PrEP have been reimbursed by statutory health insurance in Germany. While a considerable fraction of PrEP-eligible individuals receives PrEP, coverage is inhomogeneous across Germany. This study aims to identify potential barriers associated with PrEP non-use.

**Methods:** Based on the PrApp online cross-sectional study, we analyzed 1,027 PrEP users and 431 non-PrEP users. A PrEP indication was assumed for cis-MSM with an STI diagnosis (12 months), ≥ 2 sex partners or sexualized drug use (6 months). Characteristics between PrEP users and PrEP non-users were compared descriptively and using multivariable logistic regression.

**Results:** Non-PrEP users were more likely to be aged 18-29 years old (*P* < 0.05) and to use drugs during sex (*P* < 0.01). The highest PrEP prescriber density (*P* < 0.01) was associated with PrEP use. Fear of side effects (54.5%) was the most common barrier. Persons with sexualized drug use were more likely to report daily PrEP use as a barrier (34.3% vs. 16.9%, *P* < 0.01, adjusted *P* < 0.05).

**Conclusions:** Our analyses indicated structural barriers to PrEP use in federal states with a low HIV-specialists density. For those engaging in sexualized drug use, daily PrEP uptake could potentially be overcome by long-acting PrEP.

## Introduction

HIV remains a relevant public health threat, with globally 1.3 million new infections, 39.9 million people living with HIV (PWHIV) and 630,000 HIV-related deaths in 2023 [1]. In 2023, there were more than 96,700 PWHIV in Germany, by mode of transmission, 57,000 cases were transmitted through sex between men, 11,700 through heterosexual contact, and 9,000 through intravenous drug use [2]. Men who have sex with men (MSM) denote the most HIV affected group in Germany, accounting for 54.5% (1,200) of the estimated 2,200 new HIV infections in 2023 [2].

While antiretroviral drugs are successfully used for the management of an infection, immediate and successful antiretroviral treatment of PWHIV (TasP) reduces patients’ viral loads below the limit of detection and therefore prevents further sexual transmission [3]. Another important public health strategy is HIV pre-exposure prophylaxis (PrEP), where antivirals are taken before a potential HIV exposure. PrEP with daily oral emtricitabine/tenofovir disoproxil (FTC/TDF) was approved by the U.S. Food and Drug Administration (FDA) in 2012, and in 2016 by the European Medicines Agency (EMA) [4]. Several clinical trials demonstrated that daily oral PrEP can effectively protect individuals at risk from an HIV infection [5–8], and two trials demonstrated prophylactic efficacy of oral PrEP in MSM, when taken on demand [9, 10]. More recently, long-acting antiretrovirals, such as bimonthly cabotegravir, or 6-montly lenacapavir injections demonstrated HIV risk reductions comparable to those of oral PrEP in fully adherent MSM [6, 7]. While long-acting PrEP may be more convenient to use for some individuals struggling with daily adherence, to date, oral TDF/FTC PrEP is widely available as a generic [11]. In fact, over 99% of the estimated ≈ 8 million PrEP initiations worldwide are with oral TDF/XTC [12]. Oral TDF/XTC-based PrEP is available in all European Union member states, but access varies considerably: For example, in France any primary care physician can prescribe PrEP as of 2021 and the costs of PrEP are reimbursed by public health insurance [13]. In the Netherlands, PrEP can be prescribed by general practitioners, but individuals need to pay for the medicine. In Germany, PrEP can be prescribed by licensed specialists to those with substantial HIV risk. Since September 2019, the costs of PrEP have been reimbursed by statutory health insurance (SHI) [14, 15].

The estimated number of PrEP users in Germany was 15,600 – 21,600 MSM by June 2020, albeit with large regional differences and unsatisfied needs in rural areas [16]. These regional differences were also reflected in the estimated PrEP coverage, which was 24% (CI: 24–25%) by the end of 2020 [17]. The projected number of PrEP users was ≈ 50,000 by December 2025 [17] in Germany (population 84 millions), which stands in contrast to the estimated 80,000 PrEP initiations in France (population 68 millions) [12], which has the lowest barrier to PrEP access among European Union member states, albeit not reaching all individuals in need of PrEP [18]. To increase PrEP coverage, it is thus necessary to understand financial, structural, societal and behavioral barriers that may explain why PrEP-eligible MSM are not taking PrEP. In this work, we analyzed data from an anonymized online-survey among MSM PrEP users and -non-users to identify potential barriers to PrEP uptake in non-PrEP users with a PrEP indication in Germany.

## Methods

### Study design

The PrApp study is a cross-sectional study on PrEP uptake among MSM in Germany [19]. Participants were recruited through MSM dating apps (Grindr, Planetromeo and Hornet), anonymous testing checkpoints and a community website (https://prepjetzt.de/). In addition, participants were encouraged to recruit peers. Consent was required to complete the anonymous online survey, which was offered in multiple languages and included questions on sexual behavior, sexually transmitted infections (STIs) and reasons for not using PrEP. Data from two survey waves (February - May 2020 and November - December 2020) were used in this analysis.

### Participant selection

We included all cis-male participants who completed the survey and either were not using PrEP or were currently using PrEP. Moreover, participants from the last survey wave were excluded if they had participated in a previous wave to avoid duplicates. Concerning the non-PrEP users, only those who confirmed being HIV-negative were included. In order to identify those with a substantial HIV risk (= PrEP indication) the participants were filtered using criteria based on the German-Austrian PrEP guidelines [20]. A PrEP indication for non-PrEP users existed if at least one of the following criteria applied: sexualized drug use in the last six months, STI diagnosis (syphilis, gonorrhea, chlamydia or hepatitis C) in the last twelve months, or two or more sexual partners in the last six months while simultaneously having a low condom use (0-50%). The same criteria, except for low condom use, were applied to PrEP users. Low condom use was excluded from the criteria for PrEP indication for PrEP users since previous studies reported infrequent condom use among PrEP users [19, 21].

### Outcomes and covariates

The outcomes of this analysis were variables associated with non-PrEP uptake and reasons for not using PrEP among those with a PrEP indication.

A complete list of survey questions can be found in the Appendix (Suppl. Table S1). The categories ’I don’t know’ and ’Prefer not to say’ were removed from the variables and therefore treated as missing values. In terms of demographic characteristics, age groups were identified as 18-29, 30-39, 40-49 and 50-80 years. We grouped gender as cis-male (gender identity and sex assigned at birth were male), cis-female (gender identity and sex assigned at birth were female) and gender diverse (gender identity was trans, non-binary or inter, or gender identity was different from sex assigned at birth). Furthermore, country of origin was summarized as ’Germany’ and ’Outside Germany’. School leaving qualification was defined as ’No school leaving qualification’, ’Secondary school qualification (class 8 to 9)’, ’Secondary school qualification / O-Levels (class 10)’, ’A-Levels (class 12 or 13)’. Satisfaction with sex life was grouped into ’Content’ (combining happy and very happy), ’Discontent’ (combining unhappy and very unhappy) and ’Sex doesn’t matter right now’. We grouped the number of anal or vaginal sex partners within the last six months into 0, 1, 2-3, 4-5, 6-10, 11-20 and >20. The frequency of condom use for anal or vaginal sex was categorized into 0%, 25%, 50%, 75% and >95%. Regarding the number of sexual intercourse in the last month the categories were defined as 0, 1-4x, 5-8x, 9-12x, >12x. Gender of sex partners was categorized as male, female and non-binary with each category as an individual variable, allowing for multiple answers. Sexualized drug use in the last six months, payment for sex in the last six months and STI symptoms in the last 12 months had the categories ’Yes’ and ’No’. Injection of drugs during sex was categorized into ’No’, ’Yes, but not in the last 6 months’, ’Yes, 1-3x in the last 6 months’ and ’Yes, more than 3x in the last 6 months’. Regarding having had a positive STI test in your life this had the categories ’Yes’ and ’No’ for syphilis, gonorrhea, chlamydia, hepatitis C and never having had an STI. The number of STI diagnosis in the last 12 months (separately for syphilis, gonorrhea, chlamydia, hepatitis C) was grouped into 0, 1 and ≥2. The grouping is summarized in Suppl. Table S2.

The federal states of residence were encoded as the number of HIV-specialists per 10,000 gay men for each federal state and then grouped into 0, 1-2, 3-5, 6-9 and 10-13 as HIV-specialists density by federal state as described elsewhere [16]. The number of HIV-specialists is based on the website of the German association of physicians in private practice providing HIV-care “Deutsche Arbeitsgemeinschaft niedergelassener Arzte in der Versorgung HIV-Infizierter e. V. ” (dagnä) (30.06.2020) [22] and the estimate of the gay population in each federal state from the European MSM Internet Survey 2017 (EMIS) [23].

Moreover, the first three digits of the postcode were used to determine whether the participant lived in a rural or urban area based on data from the federal institute for research on building, urban affairs and spatial development “Bundesinstitut für Bau-, Stadt-und Raumforschung” (BBSR) [24]. The postcode was utilized to find the matching administrative seat in the BBSR data and then further spatial information was derived from this. For some postal codes, the matching administrative unit had to be determined manually, as an administrative seat included several postcodes. Based on the monthly household net income the monthly net equivalent income was calculated by dividing the mean of the monthly household net income category by the square root of the number of persons in the household [25]. For the highest category the population-weighted median of persons with a household net income over 5000€ was used, which was 6260€ (SOEP 2018) [26]. The monthly net equivalent income was categorized into <1,000€, 1,000– <2,000€, 2,000–<3,000€, 3,000–<4,000€, 4,000–<5,000€, and ≥5,000€.

### Statistical methods

Categorical variables are shown as absolute numbers and proportions. In univariate analyses, differences in the distribution of reasons for not using PrEP were analyzed using the Pearson’s chi-squared test and for low frequencies with Fisher’s exact test with an alpha level of 0.05. The reasons for non-PrEP uptake were analyzed in relation to PrEP indication and HIV-specialist density. *P*-values were adjusted with the Benjamini-Hochberg procedure to correct for multiple testing.

For the multivariable analysis the variables were one-hot encoded, meaning each category was encoded as a new binary variable, except for the most frequent category which was used as reference to avoid multicollinearity. Furthermore, missing values were imputed as 0 (i.e. set to the reference category). We employed a multivariable logistic regression model with elastic net regularization and tuned the hyper-parameters (mixing parameter λ and the regularization strength α). First the optimal regularization strength was determined by 15-fold stratified repeated cross-validation for a fixed mixing parameter λ∈[0.1, 0.2, 0.3, 0.4, 0.5, 0.6, 0.7, 0.8, 0.9, 1.0] and the optimal regularization strength was then chosen based on the elbow point (maximized AUC score and minimized number of non-zero coefficients). Next, for each mixing parameter λ a regression model was fitted with the selected optimal regularization strength. The model with the lowest number of non-zero coefficients and lowest Akaike-Information-Criterion (AIC) score was chosen. Finally, the final selected logistic regression model was used in a stratified bootstrap with 10,000 samples with replacement, resulting in the bootstrapped regression coefficients, which were then used to calculate the 95% percentile confidence interval and to determine the single sided bootstrap-based *P*-values. In a sensitivity analysis, the multivariable analysis was repeated including missing values as a separate category (instead of imputing them as the reference category).

### Data availability

The PrApp study was approved by the ethics commission of the Berlin Chamber of Physicians (Ref: Eth-14/18). All participants had to provide informed consent through the survey website before starting the survey. The raw survey data are not publicly available to preserve individuals’ privacy under the European General Data Protection Regulation. An example minimum dataset is available through github (https://github.com/KleistLab/PrAppData).

### Code availability

All analyses were performed in Python version 3.13.1 using custom codes. All codes are available through github (https://github.com/KleistLab/PrAppData).

## Results

### Participants

Overall, 566,933 visitors were directed to the study website in the two survey waves, with 3,623 starting the survey and 3,581 accepting the participation. 62 participants, who did not specify their PrEP uptake, were excluded. The 3,519 participants (first wave: 2,625, second wave: 894) were further selected for study completion (2,902) and study completion and being cis-male (2,828). Of the latter, 1,628 participants were not using PrEP and 1,104 were currently using PrEP. 96 Participants who answered that they were former PrEP users were excluded from the analysis. 54 participants from the second wave, who had already participated in a previous wave, were identified as duplicates and removed from analysis. Further, 197 non-PrEP users who didn’t confirm to be HIV-negative were excluded. The remaining 1,420 non-PrEP and 1,061 current PrEP users were filtered according to whether they fulfilled the criteria for PrEP indication. In the end, there were 431 non-PrEP users with a PrEP indication and 1,027 current PrEP users with a PrEP indication (Fig. 1).

**Fig. 1:**
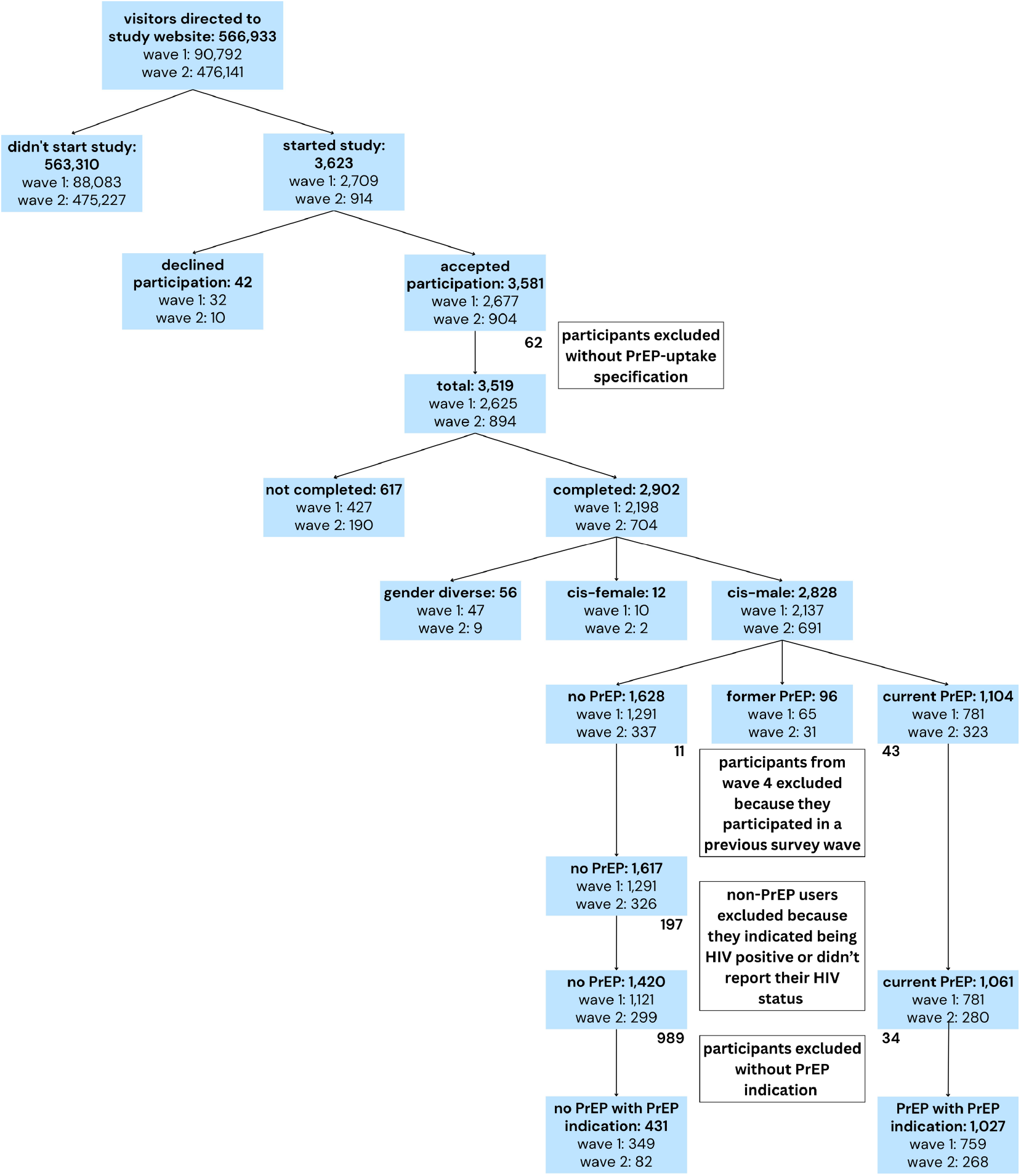
PrApp participant selection for analysis of factors associated with PrEP use. The two survey waves were conducted February - May 2020 and November - December 2020.

To understand if the data is representative across Germany or has a spatial bias, we calculated the probability of participation in the PrApp study for each federal state based on the estimated gay population, as shown in Table 1. The participation probability ranged from 0.12% (Schleswig-Holstein) to 0.47% (Berlin), indicating a comparable probability of participation between the federal states.

**Table 1:** Probability of participating in PrApp study by federal state, CI: Wilson confidence interval

| Federal state | Estimated gay population<br>(EMIS 2017), n | PrApp Participants with<br>PrEP indication, n | Probability of participation,<br>% (95% CI) |
| --- | --- | --- | --- |
| Baden-Württemberg | 44,651 | 115 | 0.26% (0.21-0.31%) |
| Bavaria | 54,061 | 212 | 0.39% (0.34-0.45%) |
| Berlin | 59,394 | 282 | 0.47% (0.42-0.53%) |
| Brandenburg | 8,395 | 20 | 0.24% (0.15-0.37%) |
| Bremen | 3,930 | 9 | 0.23% (0.12-0.43%) |
| Hamburg | 17,713 | 78 | 0.44% (0.35-0.55%) |
| Hesse | 31,422 | 134 | 0.43% (0.36-0.50%) |
| Mecklenburg Western Pomerania | 6,329 | 19 | 0.30% (0.19-0.47%) |
| Lower Saxony | 28,175 | 79 | 0.28% (0.23-0.35%) |
| North Rhine-Westphalia | 79,376 | 301 | 0.38% (0.34-0.42%) |
| Rhineland-Palatinate | 13,156 | 57 | 0.43% (0.33-0.56%) |
| Saarland | 4,520 | 12 | 0.27% (0.15-0.46%) |
| Saxony | 20,499 | 57 | 0.28% (0.21-0.36%) |
| Saxony-Anhalt | 7,454 | 15 | 0.20% (0.12-0.33%) |
| Schleswig-Holstein | 13,395 | 16 | 0.12% (0.07-0.19%) |
| Thuringia | 6,956 | 12 | 0.17% (0.10-0.30%) |

Current PrEP users had a median age of 38 years (IQR: 31-45), while non-PrEP users had a lower median age of 35 years (IQR: 28-43.5), depicted in Table 2. The majority of the PrApp participants with a PrEP indication were born in Germany (81.2%). On the one hand, a higher proportion of non-PrEP users indicated a net equivalent income of <1000€ (17.2% vs. 7.7%), very consistent condom use (22.5% vs. 8.4%), 2-3 sex partners (31.1% vs. 12.0%), as well as sexualized substance use (30.2% vs. 19.6%). On the other hand, more PrEP users were living in a federal state with a high HIV-specialists density (28.0% vs. 18.18%), did not use condoms (31.4% vs. 14.4%) and had >20 sex partners (27.8% vs. 10.9%).

**Table 2:** Baseline characteristics of PrApp participants with PrEP indication and the PrEP indication filter criteria, stratified by PrEP and non-PrEP users.

|  | Participants, n (%) | PrEP users, n (%) | Non-PrEP users, n (%) |
| --- | --- | --- | --- |
| **Total (n)** | 1,458 | 1,027 | 431 |
| **Age (years)** |  |  |  |
| Median (IQR) | 37 (30-45) | 38 (31-45) | 35 (28-43.5) |
| 18-29, n (%) | 337 (23.1%) | 201 (19.6%) | 136 (31.6%) |
| 30-39, n (%) | 490 (33.6%) | 358 (34.9%) | 132 (30.6%) |
| 40-49, n (%) | 403 (27.6%) | 306 (29.8%) | 97 (22.5%) |
| 50-80, n (%) | 228 (15.6%) | 162 (15.8%) | 66 (15.3%) |
| Missing | 0 (0.0%) | 0 (0.0%) | 0 (0.0%) |
| **Country of origin, n (%)** |  |  |  |
| Germany | 1,184 (81.2%) | 847 (82.5%) | 337 (78.2%) |
| Outside Germany | 274 (18.8%) | 180 (17.5%) | 94 (21.8%) |
| Missing | 0 (0.0%) | 0 (0.0%) | 0 (0.0%) |
| **Monthly net equivalent income, n (%)** |  |  |  |
| <1,000€ | 153 (10.5%) | 79 (7.7%) | 74 (17.2%) |
| 1,000 – 1,999€ | 360 (24.7%) | 234 (22.8%) | 126 (29.2%) |
| 2,000 – 2,999€ | 414 (28.4%) | 294 (28.6%) | 120 (27.8%) |
| 3,000 – 3,999€ | 238 (16.3%) | 189 (18.4%) | 49 (11.4%) |
| 4,000 – 4,999€ | 176 (12.1%) | 143 (13.9%) | 33 (7.7%) |
| ≥5,000€ | 54 (3.7%) | 44 (4.3%) | 10 (2.3%) |
| Missing | 63 (4.3%) | 44 (4.3%) | 19 (4.4%) |
| **HIV-specialists density, n (%)** |  |  |  |
| 0 | 16 (1.1%) | 12 (1.2%) | 4 (0.9%) |
| 1-2 | 123 (8.4%) | 81 (7.9%) | 42 (9.7%) |
| 3-5 | 360 (24.7%) | 255 (24.8%) | 105 (24.4%) |
| 6-9 | 550 (37.7%) | 369 (35.9%) | 181 (42.0%) |
| 10-13 | 369 (25.3%) | 288 (28.0%) | 81 (18.8%) |
| Missing | 40 (2.7%) | 22 (2.1%) | 18 (4.2%) |
| **Condom use, n (%)** |  |  |  |
| 0% | 384 (26.3%) | 322 (31.4%) | 62 (14.4%) |
| 25% | 452 (31.0%) | 339 (33.0%) | 113 (26.2%) |
| 50% | 255 (17.5%) | 154 (15.0%) | 101 (23.4%) |
| 75% | 161 (11.0%) | 106 (10.3%) | 55 (12.8%) |
| >95% | 183 (12.6%) | 86 (8.4%) | 97 (22.5%) |
| Missing | 23 (1.6%) | 20 (1.9%) | 3 (0.7%) |
| **Sex partners in the last 6 months, n (%)** |  |  |  |
| 0 | 10 (0.7%) | 2 (0.2%) | 8 (1.9%) |
| 1 | 24 (1.6%) | 3 (0.3%) | 21 (4.9%) |
| 2-3 | 257 (17.6%) | 123 (12.0%) | 134 (31.1%) |
| 4-5 | 265 (18.2%) | 164 (16.0%) | 101 (23.4%) |
| 6-10 | 325 (22.3%) | 242 (23.6%) | 83 (19.3%) |
| 11-20 | 228 (15.6%) | 193 (18.8%) | 35 (8.1%) |
| >20 | 333 (22.8%) | 286 (27.8%) | 47 (10.9%) |
| Missing | 16 (1.1%) | 14 (1.4%) | 2 (0.5%) |
| **Positive STI diagnosis in the last 12 months, n (%)** |  |  |  |
| Yes | 549 (37.7%) | 413 (40.2%) | 136 (31.6%) |
| No | 324 (22.2%) | 231 (22.5%) | 93 (21.6%) |
| Missing | 585 (40.1%) | 383 (37.3%) | 202 (46.9%) |
| **Sexualized substance use in the last 6 months, n (%)** |  |  |  |
| Yes | 331 (22.7%) | 201 (19.6%) | 130 (30.2%) |
| No | 1,097 (75.2%) | 808 (78.7%) | 289 (67.1%) |
| Missing | 30 (2.1%) | 18 (1.8%) | 12 (2.8%) |

The highest HIV-specialists density in 2020 could be found in the city states Berlin (10.27 HIV-specialists per 10,000 gay men), Hamburg (10.73 HIV-specialists per 10,000 gay men) and Bremen (12.73 HIV-specialists per 10,000 gay men), while the lowest HIV-specialists densities were observed in Mecklenburg Western Pomerania, Brandenburg and Schleswig-Holstein (Fig. 2). There was a tendency towards a lower HIV-specialists density in the east German federal states. The proportion of PrEP users among PrApp participants for each federal state varied between 0.40 (Saxony-Anhalt) and 0.89 (Bremen), depicted in Table 3.

**Table 3:** PrEP user proportion among PrApp participants with PrEP indication.

| Federal state | PrApp Participants with<br>PrEP indication, n | PrEP users with<br>PrEP indication, n | PrEP users density among<br>participants |
| --- | --- | --- | --- |
| Baden-Württemberg | 115 | 72 | 0.63 |
| Bavaria | 212 | 155 | 0.73 |
| Berlin | 282 | 218 | 0.77 |
| Brandenburg | 20 | 12 | 0.60 |
| Bremen | 9 | 8 | 0.89 |
| Hamburg | 78 | 62 | 0.79 |
| Hesse | 134 | 99 | 0.74 |
| Mecklenburg Western Pomerania | 19 | 14 | 0.74 |
| Lower Saxony | 79 | 56 | 0.71 |
| North Rhine-Westphalia | 301 | 198 | 0.66 |
| Rhineland-Palatinate | 57 | 43 | 0.75 |
| Saarland | 12 | 8 | 0.67 |
| Saxony | 57 | 36 | 0.63 |
| Saxony-Anhalt | 15 | 6 | 0.40 |
| Schleswig-Holstein | 16 | 12 | 0.75 |
| Thuringia | 12 | 6 | 0.50 |

**Fig. 2:**
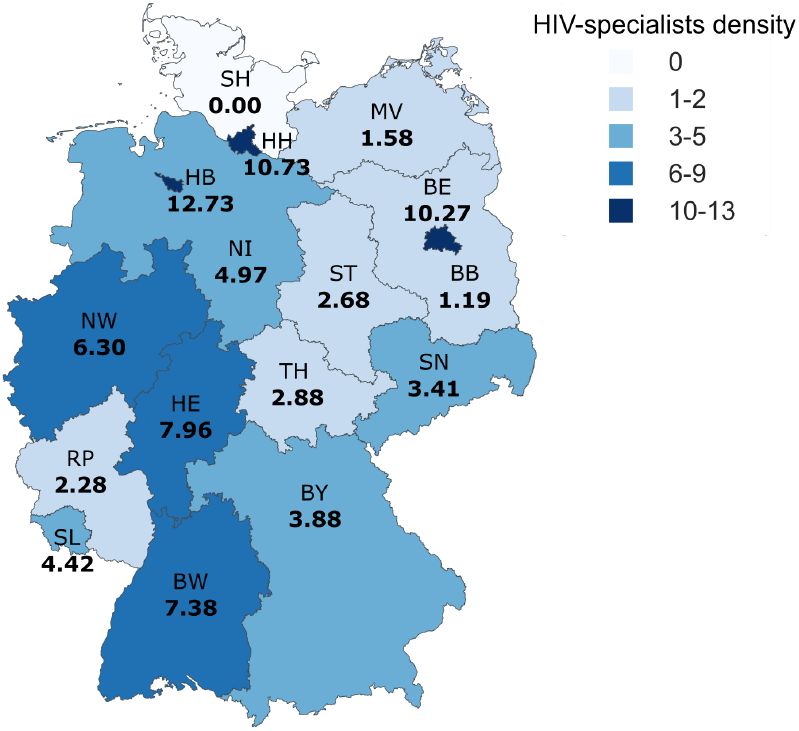
HIV-specialists density by federal state. The number of HIV specialists per 10,000 gay men is reported in bold. The federal states are abbreviated as follows: BW = Baden-Württemberg, BY = Bavaria, BE = Berlin, BB = Brandenburg, HB = Bremen, HH = Hamburg, HE = Hesse, MV = Mecklenburg Western Pomerania, NI = Lower Saxony, NW = North Rhine-Westphalia, RP = Rhineland Palatinate, SL = Saarland, SN = Saxony, ST = Saxony-Anhalt, SH = Schleswig-Holstein, TH = Thuringia

### Variables associated with not using PrEP

The multivariable analysis resulted in 13 variables associated with using or not using PrEP (*P*-value < 0.05), which are depicted with their strata in Fig. 3. A complete list of all variables and their regression coefficients with corresponding 95% CIs and *P-*values are depicted in Suppl. Table S3. Regarding sexual behavior the analysis showed that a lower number of sex partners in last six months (1-5 sex partner), as well as sexualized substance use in the last six months was associated with non-PrEP users, while a higher number of sex partners in the last six months (11-20 sex partner) and having had a chlamydia diagnosis in the past was more pronounced in PrEP users. This could be explained by the fact that PrEP users are much more frequently screened for chlamydia than non-PrEP users because asymptomatic STI screening for PrEP users is reimbursed. Non-PrEP users were more likely 18-29 years old and also had female sexual partners more frequently (10.7% vs. 4.3%, *P* = 0.056) than PrEP users. Additionally, non-PrEP users were more likely discontent with their sex life. In terms of structural barriers a higher HIV-specialists density in the federal state of residence was associated with PrEP use, while a low monthly net equivalent income of 1-<1000€ was associated with non-PrEP use.

**Fig. 3:**
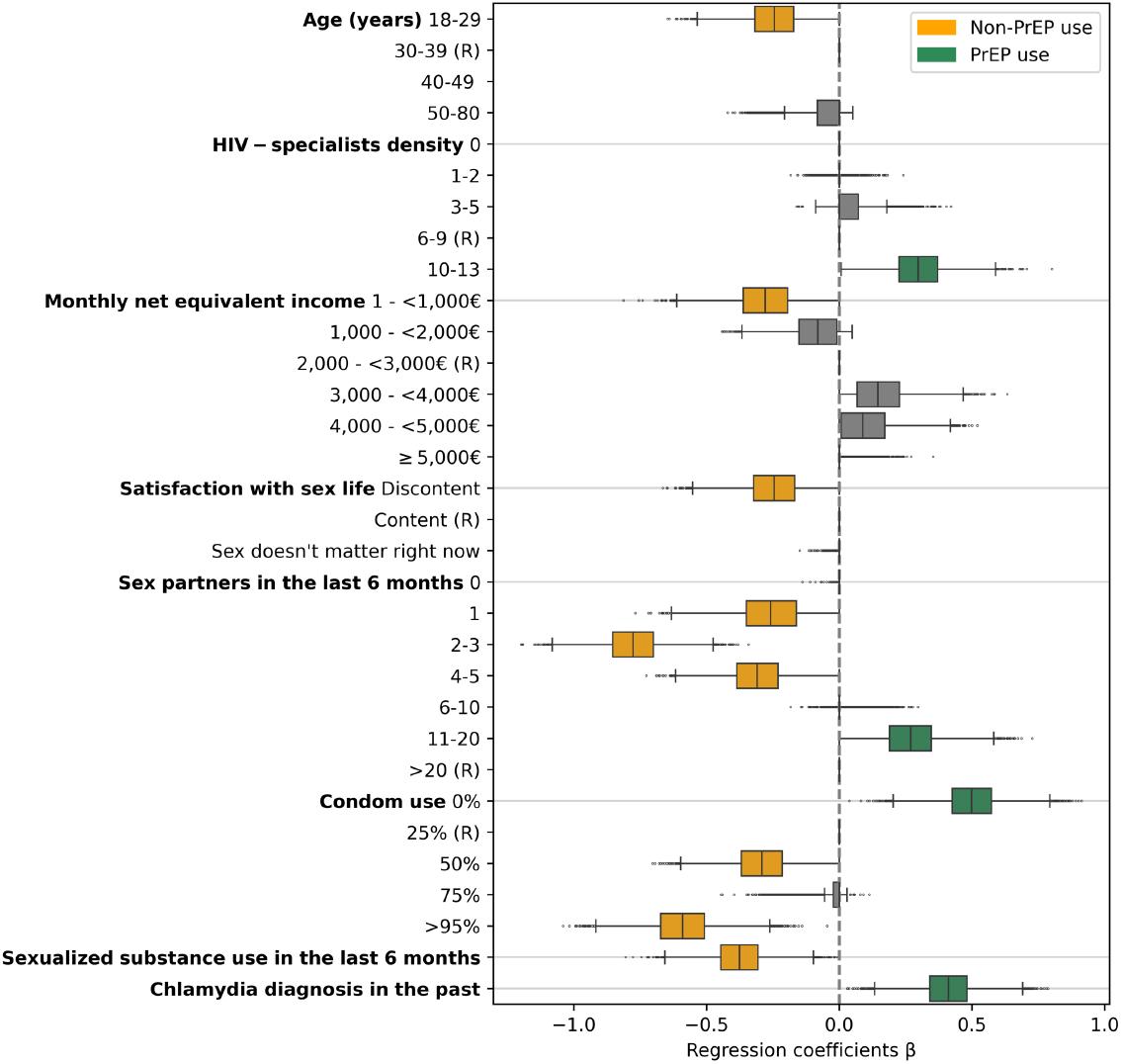
Distribution of bootstrapped regression coefficients for the significant variables (in orange and green) with *P*-value < 0.05 and their strata (in grey), (R) indicates the reference category, which is just depicted for completeness (no coefficients estimated). For some categories, the interquartile ranges of bootstrapped coefficient estimates is zero with no boxplot depicted or only outliers as dots.

The results of the sensitivity analysis, the repeated multivariable analysis including missing values as a separate category, can be found in the Appendix (Suppl. Table S4).

### Reasons for not taking PrEP

To identify potential barriers to PrEP uptake, non-PrEP users with a PrEP indication were asked for their reasons for not taking PrEP, allowing multiple answers. The most common reason for not taking PrEP in non-PrEP users with a PrEP indication was the fear of side effects (54.5%), followed by the related effort of doctor visits and regular testing (34.3%) as depicted in Table 4. The reasons for not taking PrEP were stratified by PrEP indication. We observed differences between non-PrEP users with a PrEP indication due to their sexual behavior and/or STI history and non-PrEP users with a PrEP indication due to sexualized substance use, but due to the effect sizes these were mostly not significant at the *P* < 0.05 level. In non-PrEP users with sexualized substance use, the necessity for daily intake of PrEP was identified as a barrier (34.3% vs. 16.9%, non-adjusted *P* = 0.002, adjusted *P* = 0.019), shown in Table 4.

**Table 4:** Reasons for not taking PrEP in non-PrEP users with PrEP indication, stratified by PrEP indication subgroups, multiple answers allowed (*P*-value was calculated with Chi-squared test, for frequency less than five with Fisher’s exact test (*), for multiple testing correction Benjamini-Hochberg was used).

| Reasons for not taking PrEP | All participants<br>(n = 431) | Both<br>(n = 63) | Sexual<br>behavior / STI<br>(n = 301) | Sexualized<br>substance use<br>(n = 67) | $P$-value | adjusted<br>$P$-value |
| --- | --- | --- | --- | --- | --- | --- |
| Personal HIV risk too low | 110 (25.5%) | 13 (20.6%) | 75 (24.9%) | 22 (32.8%) | 0.239 | 0.478 |
| No doctor prescribing PrEP | 88 (20.4%) | 19 (30.2%) | 55 (18.3%) | 14 (20.9%) | 0.746 | 0.994 |
| Effort: doctor visits + regular tests | 148 (34.3%) | 24 (38.1%) | 109 (36.2%) | 15 (22.4%) | 0.043 | 0.164 |
| Not wanting to discuss sex life with doctor | 116 (26.9%) | 19 (30.2%) | 83 (27.6%) | 14 (20.9%) | 0.333 | 0.532 |
| Negative reactions of sexual partners/others | 33 (7.7%) | 7 (11.1%) | 25 (8.3%) | 1 (1.5%) | * 0.062 | 0.164 |
| Fear of side effects | 235 (54.5%) | 27 (42.9%) | 171 (56.8%) | 37 (55.2%) | 0.920 | 1.000 |
| Daily pill too strenuous | 89 (20.6%) | 15 (23.8%) | 51 (16.9%) | 23 (34.3%) | 0.002 | 0.019 |
| Medical reasons | 4 (0.9%) | 0 (0%) | 4 (1.3%) | 0 (0%) | * 1.000 | 1.000 |

Perceiving the personal HIV risk as too low (32.8% vs. 24.9%, non-adjusted *P* = 0.239, adjusted *P* = 0.478) was more frequently associated with non-PrEP use with a PrEP indication due to sexualized substance use. Furthermore, the fear of negative reactions of sex partners or others (27.6% vs. 20.9%, non-adjusted *P* = 0.062, adjusted *P* = 0.164) as well as the previously mentioned effort (36.2% vs. 22.4%, non-adjusted *P* = 0.043, adjusted *P* = 0.164) were more pronounced barriers in non-PrEP users who got their PrEP indication due to their sexual behavior or STI history.

Additionally, the reasons for not taking PrEP were stratified by the HIV-specialists density in the federal state of residence (Table 5). A low HIV-specialists density was defined as 0-9 HIV specialists per 10,000 gay men and a high HIV-specialists density referred to 10-13 HIV specialists per 10,000 gay men. Non-PrEP users living in a federal state with a low HIV-specialists density were more likely to not want to discuss their sex life with a doctor (29.5% vs. 16.0%, non-adjusted *P* = 0.021, adjusted *P* = 0.083). Moreover, non-PrEP users living in a federal state with a high HIV-specialists density were associated with the barriers fear of side effects (67.9% vs. 51.8%, non-adjusted *P* = 0.013, adjusted *P* = 0.083) and perceiving their HIV-risk as not high enough (35.8% vs. 24.1%, non-adjusted *P* = 0.045, adjusted *P* = 0.121).

**Table 5:** Reasons for not taking PrEP in Non-PrEP users with PrEP indication, stratified by HIV-specialists density (Low: 0-9, High: 10-13), multiple answers allowed (*P*-value was calculated with Chi-squared test, for frequency less than five with Fisher’s exact test (*), for multiple testing correction Benjamini-Hochberg was used).

| Reasons for not taking PrEP | All participants<br>(n = 431) | Low HIV-specialists<br>density (n = 332) | High HIV-specialists<br>density (n = 81) | $P$-value | adjusted<br>$P$-value |
| --- | --- | --- | --- | --- | --- |
| Personal HIV risk too low | 110 (25.5%) | 80 (24.1%) | 29 (35.8%) | 0.045 | 0.121 |
| No doctor prescribing PrEP | 88 (20.4%) | 70 (21.1%) | 12 (14.8%) | 0.266 | 0.306 |
| Effort: doctor visits + regular tests | 148 (34.3%) | 122 (36.7%) | 23 (28.4%) | 0.200 | 0.306 |
| Not wanting to discuss sex life with doctor | 116 (26.9%) | 98 (29.5%) | 13 (16.0%) | 0.021 | 0.083 |
| Negative reactions of sexual partners/others | 33 (7.7%) | 27 (8.1%) | 3 (3.7%) | 0.255 | 0.306 |
| Fear of side effects | 235 (54.5%) | 172 (51.8%) | 55 (67.9%) | 0.013 | 0.083 |
| Daily pill too strenuous | 89 (20.6%) | 65 (19.6%) | 21 (25.9%) | 0.267 | 0.306 |
| Medical reasons | 4 (0.9%) | 3 (0.9%) | 1 (1.2%) | * 0.584 | 0.584 |

## Discussion

We analyzed potential barriers to PrEP uptake by identifying differences between PrEP and non-PrEP users with a PrEP indication.

PrEP users were more likely to live in a federal state with a high HIV-specialists density, while non-PrEP users were more often younger, had a lower monthly net equivalent income, were more likely to have fewer sexual partners, and a more persistent condom use. Among non-PrEP users with a PrEP indication due to their sexualized substance use 34.3% indicated the burden of a daily pill as a barrier to PrEP uptake. The following observed differences were at a sub-critical statistical level and further evaluation by larger cohorts would be warranted: Non-PrEP users with a PrEP indication, due to their sexual behaviour and/or STI history, were more likely to name the related effort with doctor visits and regular testing (36.2%) as a barrier. Notably, non-PrEP users living in a federal state with a low HIV-specialists density reported structural reasons for not taking PrEP, such as not wanting to discuss their sex life with their doctor (29.5%). However, for non-PrEP users living in federal states with a high HIV-specialists the barriers were more individual, namely the fear of side effects (67.9%) and the perceived personal HIV risk as too low (35.8%).

Overall, the most common reason for not taking PrEP (54.5%) was the fear of side effects. Concerns about side effects as a barrier to PrEP uptake have been found in multiple studies [27–30]. Notably, the fear of long-term side effects as a reason for PrEP discontinuation was identified in former PrEP users [31]. While TDF may decreases bone mineral density [32], a systematic review on oral PrEP with TDF/FTC or TDF detected no increased risk of bone fracture, and while they identified an association with a slightly elevated risk of gastrointestinal and renal adverse events, most adverse events were minor and reversible [33]. By educating individuals who are eligible to use PrEP on the advantages and disadvantages of PrEP, including potential side effects, the fear of side effects could be addressed and personal decisions balancing the benefits and potential disadvantages of oral PrEP may be better informed.

Our study indicating a lower PrEP uptake among young MSM (18-29 years; β = 0.24, 95% CI -0.46 – -0.03, *P* = 0.013) is supported by the findings of a meta-analysis on PrEP use among PrEP-eligible MSM, which found a higher percentage of current PrEP user in studies with a median age ≥ 30 years than in studies with a median age *<* 30 years (33.1% [95% CI 23.7-44.0; PI 12.9-62.2] vs. 16.3% [95% CI 10.9-23.6; PI 5.4-40.1], *P* = 0.0053) [34]. Young adults are at greater risk of HIV acquisition, with 43% of recent HIV infections among individuals of age 18-25 in Germany between 2008 and 2014 [35]. Consequently, it is important to develop effective public health strategies that target young MSM. Interestingly, a Dutch study from 2001 [36] found that young MSM are more likely to contract HIV from their main sex partner than from casual sex partner, indicating among other things the need for relationship-level HIV prevention strategies for young MSM. However, this finding dated to the time before TasP, and consequently the risk of contracting HIV have changed considerably, as well as perceived risks [37]. Several studies indicated a high interest in long-acting injectable PrEP among young MSM, suggesting long-acting PrEP might be suitable for this group [38–40].

Non-PrEP users were also more likely to have a low monthly net equivalent income of 1-<1000€ (β = -0.30, 95% CI -0.52 – -0.03, *P* = 0.014). A study among young app-using MSM in California, as well as a retrospective study based on electronic medical records of federally-qualified health centers in New York, showed an association between PrEP use and higher income [41, 42]. Since September 2019, the costs of PrEP have been covered by statutory health insurance for individuals with substantial HIV risk. Due to the coverage, PrEP should be accessible independent of income, however, our findings suggest that the economic situation still plays a role. This is supported by a study on the changes of sociodemographic profiles of PrEP users before and after PrEP reimbursement conducted at a cross-sectoral sexual health centre in Germany, which showed no significant sociodemographic differences [43]. A potential reason could be that individuals with a lower net equivalent income are less aware that PrEP costs are reimbursed by statutory health insurance, or they don’t have the time and capacity to access PrEP. The former could be addressed by a targeted information campaign. Regarding the latter, as HIV-specialists are mostly located in urban areas, transportation to visit a PrEP prescriber is already time-consuming, which could be a barrier for MSM with a low income. By making PrEP more easily accessible, for example using telemedicine [44], PrEP uptake among low income MSM could potentially be facilitated. Telemedicine for PrEP may also address structural barriers due to low HIV-specialists density.

Non-PrEP users more often had also sex with women (10.7% vs. 4.3%), but the association was weak and might have been a chance finding (β = -0.23, 95% CI -0.48 – 0.00, *P* = 0.056). Nonetheless, studies have found that bisexual men are significantly less likely to have ever taken PrEP compared to gay men, as well as an association of experience with PrEP use and a gay sexual identity [45, 46]. Furthermore, studies show that bisexual men are also less likely to get tested for HIV [45, 47]. This may indicate that bisexual MSM are less likely to go to places that offer PrEP and HIV testing, e.g., checkpoints. One possible explanation for this could be that bisexual men don’t perceive themselves as the target group for education on HIV prevention, as well as places offering related healthcare services, and rather tend to associate this with gay men. Consequently, a communication on HIV risks and prevention addressing bisexual men is important to reach all MSM in need of PrEP.

We found that among MSM with a PrEP indication, sexualized drug use was more common in non-PrEP users (β = -0.38, 95% CI -0.58 – -0.17, *P* = 0.000). In support of our findings, reduced willingness to take PrEP was moderately associated with using methylenedioxy-methamphetamine (or ecstasy), amphetamine, marijuana, mephedrone, cocaine, heroin, gamma hydroxybutrate (GHB) or ketamine [48]. However, PrEP uptake has been frequently associated with sexualized drug use [49, 50]. Studies have identified different subgroups of MSM with substance use, which were associated with different risk behaviour [51–55]. Based on the available data we cannot differentiate between different substance use patterns and their context in our data. Future studies should focus on further contextualization.

As a potential barrier we identified daily PrEP uptake for those non-PrEP users with a PrEP indication due to their sexualized drug use (34.4% vs. 16.9%, *P* = 0.002, adjusted *P* = 0.019). Notably, it is suggested that daily PrEP adherence can be effective and feasible for MSM reporting chemsex [56]. However, having an alternative to daily PrEP, such as on-demand PrEP or long-acting PrEP could be beneficial for this subgroup. Concerning structural barriers, our analyses showed a significant association between PrEP uptake and a high HIV-specialists density (β = 0.29, 95% CI 0.09 – 0.51, *P* = 0.004), suggesting that accessibility of PrEP plays an important factor for PrEP uptake. Regional differences in the number HIV-specialists are likely a barrier for accessing PrEP [16]. Importantly, it is known that MSM with their place of residence in rural areas, access PrEP in urban areas and also across federal states. Additionally, not only the distance to a doctor prescribing PrEP can be a barrier for not taking PrEP, but also the associated stigma and lack of anonymity for PrEP care in rural areas [57]. A potential barrier for non-PrEP users living in a federal state with a low HIV-specialists density was not wanting to discuss their sex life with a doctor (29.5% vs. 16.0%, non-adjusted *P* = 0.021, adjusted *P* = 0.083). These structural barriers underline the impact of the limited number health-care providers licensed to prescribe PrEP on PrEP uptake. To address this and to cover the PrEP needs of MSM not finding a PrEP prescriber due to distance and/or lack of privacy, it is important to make PrEP more accessible for example through telemedicine [44] and more PrEP prescribers.

In contrast, non-PrEP users living in federal states with a high HIV-specialists density described rather individual reasons for not taking PrEP, such as the pronounced fear of side effects (67.9% vs. 51.8%, non-adjusted *P* = 0.013, adjusted *P* = 0.083) as well as perceiving their HIV risk as too low (35.8% vs. 24.1%, non-adjusted *P* = 0.045, adjusted *P* = 0.121) despite having a PrEP indication. Recent findings found an underestimation of HIV risk among many MSM and identified low HIV-risk perception as a barrier to PrEP uptake [58–61]. This is interesting as this group of non-PrEP users theoretically should have access to PrEP, and were still not taking it. This suggests that the pure access PrEP is not sufficient, but that the communication on HIV risks and side effects is important to help individuals assess their own HIV-risk and to allow them to make an informed decision. A positive example for the impact of effective risk communication is the case decline during the 2022 mpox outbreak in MSM in Berlin [62, 63].

Non-PrEP users with a PrEP indication due to their sexual behaviour and/or a previous STI diagnosis, more often described the effort with doctor visits and regular testing (36.2% vs. 22.4%, non-adjusted *P* = 0.043, adjusted *P* = 0.164) as a reason for not taking PrEP. A recent study has demonstrated that online-mediated and 6monthly PrEP monitoring are non-inferior to in-clinic and to 3-monthly monitoring [64], which could be an approach to reduce this barrier. Importantly, non-PrEP users with an STI diagnosis in the last 12 months, have been recently in contact with health care providers, which should be used to educate them on PrEP and their own HIV risk.

A strength of our study is that it includes current PrEP users as well as non-PrEP users. This allows us to identify barriers for non-PrEP users with a PrEP indication. Notably, a limitation is that this not a causal analysis, but an investigation into potential barriers to PrEP uptake among those with a PrEP indication. Identifying these barriers allows to gain a deeper understanding on why those advised to take PrEP are not using it in times when the health insurance covers the costs of PrEP. Similarly, it is unclear how representative this study is, especially for individuals not using online resources. This is not expected to be of great impact as the use of the internet and dating apps is widely spread among MSM [65]. Furthermore, all data is self-reported, which could potentially induce a response bias. However, many participants reported for example low condom use, sexualized substance and high number of sexual partner, which suggests that potential response biases are minor.

## Conclusions

Overall, we found that the fear of side effects is still a persistent barrier to PrEP uptake. In the context of sexualized substance use, long-acting PrEP could be an alternative as the uptake of daily pill was perceived as a barrier. Furthermore, the distance to a PrEP prescriber, as well as lack of anonymity were reasons for not taking PrEP in federal states with a low HIV-specialists density, which could be addressed by telemedicine. For non-PrEP users living in federal states with a high HIV-specialists density an effective risk communication could tackle the perception of low HIV risks, as well as the fear of side effects. In summary, PrEP-eligible MSM in Germany may face different barriers to PrEP uptake, which need to be further studied.

## Acknowledgements

HW, SI and MvK acknowledge funding through the German Federal Ministry of Research, Technology and Space (BMFTR), through project “TransPrEP” (grant number 01KI2016). HW, SI and MvK acknowledge funding through an RKI-internal research funding (Sonderforschungsmittel, Strang B). The PrApp study was funded by the German Federal Ministry of Health (BMG) on the basis of a decision by the German Bundestag. The authors thank all participants of the PrApp study.

## Author contributions

UK, UM, BGB and VB designed the study. UK, SA and JH were involved in data curation. UK coordinated the study. UK and MvK conceptualized the project, guided the modelling approach, and supervised the project. SI prepared figures. HW performed the analysis and wrote the first draft with help from UK and MvK. All authors reviewed and contributed to the manuscript.

## Competing interests

The funders had no role in study design, data collection and analysis, decision to publish, or preparation of the manuscript. The authors declare that no conflicts of interest exist.

## Notes

### Competing Interest Statement

The authors have declared no competing interest.

### Author Declarations

The PrApp study was approved by the ethics commission of the Berlin Chamber of Physicians (Ref: Eth-14/18).

