## Supplementary Tables for "Analysis of potential barriers for non-PrEP users among MSM in Germany"

### **Supplementary Table S1**: Excerpt of survey questions from PrApp survey

| **Survey questions and possible answers** |
|---|
| **Are you currently taking or have you ever taken preexposure prophylaxis (PrEP) against HIV?**<br>• Yes, I am taking PrEP on a daily basis.<br>• Yes, I take it intermittently when I think I need it<br>• Yes, I used to take PrEP, but I permanently stopped<br>• No |
| **Have you participated in this survey before?** (e.g. July – October 2018 or April/May 2019)<br>• Yes<br>• No<br>• I don't remember |
| **How old are you?** |
| **What gender do you identify with?**<br>• Male<br>• Female<br>• Inter<br>• Trans male<br>• Trans female<br>• Non-binary |
| **What gender were you assigned at birth?**<br>• Male<br>• Female<br>• Intersexual |
| **In which country were you born?** |
| **What are the first three digits of the postal code of your residence in Germany?**<br>If you are not living in Germany, please indicate the first three digits of the postal code of the place where you are predominantly staying (e.g. hotel). |
| **What is the overall net income of the household you live in?**<br>With income we mean: salaries/wages, non-wage incomes, retirement pension, unemployment benefits, housing benefits, family allowances, etc. From this income, please substract all taxes, operating expenses, and social security contributions<br>• 1 to less thann 1000€<br>• 1000 to less than 2000€<br>• 2000 to less than 3000€<br>• 3000 to less than 4000€<br>• 4000 to less than 5000€<br>• 5000€ and more |
| **How many people are living in your household?**<br>• I am living alone<br>• xxx persons |
| **Did you graduate from school?**<br>• Yes<br>• No<br>• Prefer not to say |
| **What school leaving qualification do you have?**<br>• Secondary school qualification ("Hauptschulabschluss"/ "Volksschulabschluss") class 8 to 9<br>• Secondary school qualification ("Realschulabschluss") O-Levels ("Mittlere Reife") class 10<br>• Grammar school qualification ("Gymnasium" or "EOS") A-Levels ("Abitur") class 12 or 13 Technical college qualification ("Fachhochschulreife") |
| **Are you having sex with... (multiple answers)**<br>• Men<br>• Women<br>• Non-binary people |
| **With how many different partners have you had anal and/or vaginal sex within the last 6 months?**<br>• 0<br>• 1<br>• 2-3<br>• 4-5<br>• 6-10<br>• 11-20<br>• more than 20<br>• I don't know |
| **How often have you had anal and/or vaginal sex in the last month?**<br>• Not at all<br>• 1-4 times<br>• 5-8 times<br>• 9-12 times |

**Survey questions and possible answers**

- More than 12 times
- I don't know

**Have you taken any substances in the last 6 months to have more intense or longer sex?**

e.g. GHB/GBL, crystal meth, ketamine, cocaine, speed, bath salts, mephedrone, ecstasy/MDMA

- Yes
- No
- Prefer not to say

**Did you inject these substances?**

- Yes, but not within the last 6 months
- Yes, 1-3 times within the last 6 months
- Yes, more often than 3 times within the last 6 months
- No
- Prefer not to say

**Were you paid for sex within the last 6 months?**

By payment we mean the exchange of money, gifts, or favors for sex.

- Yes
- No
- Prefer not to say

**How often do you use condoms for anal / vaginal sex in periods when you are taking PrEP?**

- Always (in more than 95% of the times)
- Often (about 75% of the times)
- About half of the times (50%)
- Sometimes (about 25% of the times)
- Never
- I don't know

**How happy are you with your sex life at the moment?**

- Very happy
- Happy
- I'm not sure
- Unhappy
- Very unhappy
- Sex is not important for me at the moment

**Have you had symptoms of a sexually transmitted infection in the last 12 months?**

e.g. pain when urinating, discharge, itching/pain in the anal area

- Yes
- No
- Prefer not to say

**For which of the following STIs have you tested positive? / Which STIs have you ever been diagnosed with?**

Select all that apply:

- Syphilis
- Gonorrhea
- Chlamydia
- Hepatitis C
- None
- I don't remember

**How often have you been diagnosed with syphilis in the last 12 months?**

- 1-10
- More than 10 times

**How often have you been diagnosed with gonorrhea in the last 12 months?**

- 1-10
- More than 10 times

**How often have you been diagnosed with chlamydia in the last 12 months?**

- 1-10
- More than 10 times

**How often have you been diagnosed with hepatitis C in the last 12 months?**

- 1-10
- More than 10 times

**Why are you not using PrEP? (Multiple answers)**

- My HIV risk is not high enough.
- I am HIV-positive.
- I cannot take PrEP due to medical reasons.
- I can't find a doctor to prescribe PrEP to me.
- I don't want to talk about my sexual life with my doctor.
- The effort of regularly visiting a doctor for refills and medical tests is too high for me
- Taking a pill every day is too demanding for me.
- I am afraid of negative reactions from others / sexual partners.
- I am afraid of side effects.

**Supplementary Table S2**: Grouping of variables. (R) indicates the reference category, which was dropped in the multivariable analysis.

| Variable | Grouping |
|---|---|
| **Age** | 18–29<br>30–39 (R)<br>40–49<br>50–80 |
| **Country of origin** | Germany / Outside Germany |
| **Urban vs. rural area** | urban area (R)<br>rural area |
| **HIV-specialists density in federal state of residence** | 0<br>1-2<br>3-5<br>6-9 (R)<br>10-13 |
| **School leaving certificate** | no school leaving certificate<br>secondary school qualification (class 8 to 9)<br>secondary school qualification (class 10)<br>A-Levels (class 12 or 13) (R) |
| **Monthly net equivalent income** | <1,000€<br>1,000 – 1,999€<br>2,000 – 2,999€ (R)<br>3,000 – 3,999€<br>4,000 – 4,999€<br>≥5,000€ |
| **Satisfaction with sexlife** | content (R)<br>discontent<br>sex doesnt matter right now |
| **Sex with men** | yes / no |
| **Sex with women** | yes / no |
| **Sex with non-binary people** | yes / no |
| **Sex partners in the last 6 months** | 0<br>1<br>2-3<br>4-5<br>6-10<br>11-20<br>>20 (R) |
| **Sex frequency last month** | 0x<br>1-4x (R)<br>5-8x<br>9-12x<br>>12x |
| **Sexualized substance use in the last 6 months** | yes / no |
| **Injective sexualized substance use** | yes, but not in the last 6 months<br>yes, 1-3x in the last 6 months<br>yes, more than 3x in the last 6 months<br>no (R) |
| **Payment for sex in the last 6 months** | yes / no |
| **Condom use** | 0%<br>25% (R)<br>50%<br>75%<br>>95% |
| **STI symptoms in the last 12 months** | yes / no |
| **Syphilis diagnosis in the past** | yes / no |
| **Gonorrhea diagnosis in the past** | yes / no |
| **Chlamydia diagnosis in the past** | yes / no |
| **Hepatitis C diagnosis in the past** | yes / no |
| **No STI diagnoses in the past** | yes / no |
| **Syphilis diagnoses in the last 12 months** | 0 (R)<br>1<br>≥ 2 |
| **Gonorrhea diagnoses in the last 12 months** | 0<br>1 (R)<br>≥ 2 |
| **Chlamydia diagnoses in the last 12 months** | 0<br>1 (R)<br>≥ 2 |
| **Hepatitis C diagnoses in the last 12 months** | 0 /1 |

**Supplementary Table S3**: Comparison of PrEP and non-PrEP users with a PrEP indication, along with bootstrapped regression coefficients and *P*-values.

| | PrEP users, n (%) | Non-PrEP users, n (%) | β (95% CI) | *P*-value |
|---|---|---|---|---|
| **Total (n)** | 1,027 | 431 | | |
| **Age (years)** | | | | |
| Median (IQR) | 38 (31 - 45) | 35 (28 - 43.5) | | |
| 18–29, n (%) | 201 (19.6%) | 136 (31.6%) | -0.24 (-0.46 - -0.03) | 0.013 |
| 30–39, n (%) | 358 (34.9%) | 132 (30.6%) | 0 | |
| 40–49, n (%) | 306 (29.8%) | 97 (22.5%) | 0.07 (0.00 - 0.27) | 0.308 |
| 50–80, n (%) | 162 (15.8%) | 66 (15.3%) | -0.01 (-0.23 - 0.00) | 0.502 |
| Missing, n (%) | 0 (0%) | 0 (0%) | - | |
| **Country of origin, n (%)** | | | | |
| Germany | 847 (82.5%) | 337 (78.2%) | 0.00 (0.00 - 0.18) | 0.617 |
| Outside Germany | 180 (17.5%) | 94 (21.8%) | 0 | |
| Missing | 0 (0%) | 0 (0%) | - | |
| **Urban-rural area (based on plz), n (%)** | | | | |
| Urban area | 900 (87.6%) | 352 (81.7%) | 0 | |
| Rural area | 105 (10.2%) | 61 (14.2%) | 0.00 (-0.21 - 0.00) | 0.575 |
| Missing | 22 (2.1%) | 18 (4.2%) | - | |
| **Density of HIV-specialists in federal state of residence, n (%)** | | | | |
| 0 | 12 (1.2%) | 4 (0.9%) | 0.00 (0.00 - 0.00) | 1.000 |
| 1-2 | 81 (7.9%) | 42 (9.7%) | 0.00 (0.00 - 0.00) | 0.976 |
| 3-5 | 255 (24.8%) | 105 (24.4%) | 0.00 (0.00 - 0.21) | 0.518 |
| 6-9 | 369 (35.9%) | 181 (42.0%) | 0 | |
| 10-13 | 288 (28.0%) | 81 (18.8%) | 0.29 (0.09 - 0.51) | 0.004 |
| Missing | 22 (2.1%) | 18 (4.2%) | - | |
| **Type of school qualification, n (%)** | | | | |
| No school leaving qualification | 7 (0.7%) | 9 (2.1%) | 0.00 (0.00 - 0.00) | 0.985 |
| Secondary school qualification (class 8/9) | 49 (4.8%) | 20 (4.6%) | 0.00 (0.00 - 0.00) | 0.994 |
| Secondary school qualification (class 10) | 190 (18.5%) | 104 (24.1%) | -0.11 (-0.33 - 0.00) | 0.159 |
| A-Levels (class 12 or 13) | 776 (75.6%) | 288 (66.8%) | 0 | |
| Missing | 5 (0.5%) | 10 (2.3%) | - | |
| **Monthly net equivalent income, n (%)** | | | | |
| <1,000€ | 79 (7.7%) | 74 (17.2%) | -0.30 (-0.52 - -0.03) | 0.014 |
| 1,000 – 1,999€ | 234 (22.8%) | 126 (29.2%) | -0.09 (-0.28 - 0.00) | 0.225 |
| 2,000 – 2,999€ | 294 (28.6%) | 120 (27.8%) | 0 | |
| 3,000 – 3,999€ | 189 (18.4%) | 49 (11.4%) | 0.15 (0.00 - 0.39) | 0.103 |
| 4,000 – 4,999€ | 143 (13.9%) | 33 (7.7%) | 0.09 (0.00 - 0.33) | 0.232 |
| ≥5,000€ | 44 (4.3%) | 10 (2.3%) | 0.00 (0.00 - 0.09) | 0.885 |
| Missing | 44 (4.3%) | 19 (4.4%) | - | |
| **Sexlife, n (%)** | | | | |
| Content | 717 (69.8%) | 206 (47.8%) | 0 | |
| Discontent | 148 (14.4%) | 108 (25.1%) | -0.25 (-0.47 - -0.02) | 0.018 |
| Sex doesnt matter right now | 12 (1.2%) | 14 (3.2%) | 0.00 (0.00 - 0.00) | 0.993 |
| Missing | 150 (14.6%) | 103 (23.9%) | - | |
| **Number of sex partners within the last 6 months, n (%)** | | | | |
| 0 | 2 (0.2%) | 8 (1.9%) | 0.00 (0.00 - 0.00) | 0.996 |
| 1 | 3 (0.3%) | 21 (4.9%) | -0.27 (-0.52 - 0.00) | 0.042 |
| 2-3 | 123 (12.0%) | 134 (31.1%) | -0.78 (-0.99 - -0.55) | 0.000 |
| 4-5 | 164 (16.0%) | 101 (23.4%) | -0.32 (-0.53 - -0.08) | 0.004 |
| 6-10 | 242 (23.6%) | 83 (19.3%) | 0.00 (0.00 - 0.11) | 0.831 |
| 11-20 | 193 (18.8%) | 35 (8.1%) | 0.27 (0.04 - 0.50) | 0.011 |
| >20 | 286 (27.8%) | 47 (10.9%) | 0 | |
| Missing | 14 (1.4%) | 2 (0.5%) | - | - |
| **Condom use, n (%)** | | | | |
| 0% | 322 (31.4%) | 62 (14.4%) | 0.50 (0.29 - 0.71) | 0.000 |
| 25% | 339 (33.0%) | 113 (26.2%) | 0 | |
| 50% | 154 (15.0%) | 101 (23.4%) | -0.29 (-0.51 - -0.07) | 0.006 |
| 75% | 106 (10.3%) | 55 (12.8%) | 0.00 (-0.19 - 0.00) | 0.695 |
| >95% | 86 (8.4%) | 97 (22.5%) | -0.59 (-0.83 - -0.35) | 0.000 |
| Missing | 20 (1.9%) | 3 (0.7%) | - | - |
| **Number of sexual intercourses in the last month, n (%)** | | | | |
| 0 | 69 (6.7%) | 63 (14.6%) | -0.08 (-0.32 - 0.00) | 0.293 |
| 1-4 | 442 (43.0%) | 217 (50.3%) | 0 | |
| 5-8 | 232 (22.6%) | 68 (15.8%) | 0.03 (0.00 - 0.25) | 0.427 |
| 9-12 | 104 (10.1%) | 40 (9.3%) | 0.00 (-0.09 - 0.00) | 0.892 |
| >12 | 168 (16.4%) | 38 (8.8%) | 0.14 (0.00 – 0.38) | 0.110 |
| Missing | 12 (1.2%) | 5 (1.2%) | - | |
| **Drugs during sex in the last 6 months, n (%)** | | | | |
| Yes | 201 (19.6%) | 130 (30.2%) | -0.38 (-0.58 - -0.17) | 0.000 |
| No | 808 (78.7%) | 289 (67.1%) | 0 | |
| Missing | 18 (1.8%) | 12 (2.8%) | - | |

| | PrEP users, n (%) | Non-PrEP users, n (%) | β (95% CI) | *P*-value |
|---|---|---|---|---|
| **Injecting drugs during sex, n (%)** | | | | |
| Yes, but not in the last 6 months | 2 (0.2%) | 5 (1.2%) | 0.00 (0.00 - 0.00) | 1.000 |
| Yes, 1-3x in the last 6 months | 10 (1.0%) | 4 (0.9%) | 0.00 (0.00 - 0.00) | 1.000 |
| Yes, >3x in the last 6 months | 17 (1.7%) | 6 (1.4%) | 0.00 (0.00 - 0.00) | 1.000 |
| No | 171 (16.7%) | 114 (26.5%) | 0 | |
| Missing | 827 (80.5%) | 302 (70.1%) | - | |
| **Payment for sex in the last 6 months, n (%)** | | | | |
| Yes | 58 (5.6%) | 37 (8.6%) | 0.00 (-0.20 - 0.00) | 0.653 |
| No | 964 (93.9%) | 381 (88.4%) | 0 | |
| Missing | 5 (0.5%) | 13 (3.0%) | - | |
| **Number of syphilis diagnosis in the last 12 months, n (%)** | | | | |
| 0 | 166 (16.2%) | 52 (12.1%) | 0 | |
| 1 | 107 (10.4%) | 42 (9.7%) | 0.00 (-0.06 - 0.00) | 0.935 |
| ≥2 | 11 (1.1%) | 2 (0.5%) | 0.00 (0.00 - 0.00) | 1.000 |
| Missing | 743 (72.3%) | 335 (77.7%) | - | - |
| **Number of gonorrhoea diagnosis in the last 12 months, n (%)** | | | | |
| 0 | 173 (16.8%) | 65 (15.1%) | 0.00 (-0.18 - 0.00) | 0.663 |
| 1 | 200 (19.5%) | 56 (13.0%) | 0 | |
| ≥2 | 68 (6.6%) | 10 (2.3%) | 0.00 (0.00 - 0.13) | 0.800 |
| Missing | 586 (57.1%) | 300 (69.6%) | - | |
| **Number of chlamydia diagnosis in the last 12 months, n (%)** | | | | |
| 0 | 132 (12.9%) | 40 (9.3%) | 0.00 (-0.03 - 0.00) | 0.956 |
| 1 | 221 (21.5%) | 59 (13.7%) | 0 | |
| ≥2 | 59 (5.7%) | 2 (0.5%) | 0.10 (0.00 - 0.28) | 0.180 |
| Missing | 615 (59.9%) | 330 (76.6%) | - | |
| **Number of hepatitis C diagnosis in the last 12 months, n (%)** | | | | |
| 0 | 20 (1.9%) | 17 (3.9%) | 0 | |
| 1 | 5 (0.5%) | 1 (0.2%) | 0.00 (0.00 - 0.00) | 1.000 |
| Missing | 1,002 (97.6%) | 413 (95.8%) | - | |
| **Gender of sexual partners, n (%) (multiple responses possible)** | | | | |
| Male | 1,027 (100.0%) | 430 (99.8%) | 0.00 (0.00 - 0.00) | 1.000 |
| Female | 44 (4.3%) | 46 (10.7%) | -0.23 (-0.48 - 0.00) | 0.056 |
| Non-binary | 33 (3.2%) | 13 (3.0%) | 0.00 (0.00 - 0.00) | 0.999 |
| Missing | 0 (0%) | 0 (0%) | - | |
| **STI symptoms in the last 12 months, n (%)** | | | | |
| Yes | 352 (34.3%) | 135 (31.3%) | 0.00 (-0.14 - 0.01) | 0.726 |
| No | 673 (65.5%) | 294 (68.2%) | 0 | |
| Missing | 2 (0.2%) | 2 (0.5%) | - | |
| **Positive STI test in your life, n (%)** | | | | |
| Syphilis | 288 (28.0%) | 99 (23.0%) | 0.00 (-0.03 - 0.12) | 0.813 |
| Gonorrhoea | 444 (43.2%) | 131 (30.4%) | 0.11 (0.00 - 0.32) | 0.100 |
| Chlamydia | 420 (40.9%) | 101 (23.4%) | 0.41 (0.20 - 0.62) | 0.000 |
| Hepatitis C | 25 (2.4%) | 18 (4.2%) | 0.00 (-0.21 - 0.00) | 0.680 |
| Never | 322 (31.4%) | 177 (41.1%) | 0.00 (0.00 - 0.08) | 0.883 |

**Supplementary Table S4**: Sensitivity analysis. This includes missing values as a separate category in the multivariable analysis. Comparison of PrEP and non-PrEP users with a PrEP indication, along with bootstrapped regression coefficients and *P*-values.

| | **PrEP users, n (%)** | **Non-PrEP users, n (%)** | **β (95% CI)** | ***P*-value** |
|---|---|---|---|---|
| **Total (n)** | 1,027 | 431 | | |
| **Age (years)** | | | | |
| Median (IQR) | 38 (31 - 45) | 35 (28 - 43.5) | | |
| 18–29, n (%) | 201 (19.6%) | 136 (31.6%) | -0.24 (-0.46 - -0.03) | 0.013 |
| 30–39, n (%) | 358 (34.9%) | 132 (30.6%) | 0 | |
| 40–49, n (%) | 306 (29.8%) | 97 (22.5%) | 0.06 (0.00 - 0.26) | 0.320 |
| 50–80, n (%) | 162 (15.8%) | 66 (15.3%) | -0.03 (-0.25 - 0.00) | 0.466 |
| Missing, n (%) | 0 (0%) | 0 (0%) | 0.00 (0.00 - 0.00) | 1.000 |
| **Country of origin, n (%)** | | | | |
| Germany | 847 (82.5%) | 337 (78.2%) | 0.00 (0.00 - 0.17) | 0.662 |
| Outside Germany | 180 (17.5%) | 94 (21.8%) | 0 | |
| Missing | 0 (0%) | 0 (0%) | - | |
| **Urban-rural area (based on plz), n (%)** | | | | |
| Urban area | 900 (87.6%) | 352 (81.7%) | 0 | |
| Rural area | 105 (10.2%) | 61 (14.2%) | 0.00 (-0.22 - 0.00) | 0.540 |
| Missing | 22 (2.1%) | 18 (4.2%) | 0.00 (-0.16 - 0.00) | 0.616 |
| **Density of HIV-specialists in federal state of residence, n (%)** | | | | |
| 0 | 12 (1.2%) | 4 (0.9%) | 0.00 (0.00 - 0.00) | 1.000 |
| 1-2 | 81 (7.9%) | 42 (9.7%) | 0.00 (0.00 - 0.00) | 0.979 |
| 3-5 | 255 (24.8%) | 105 (24.4%) | 0.00 (0.00 - 0.20) | 0.551 |
| 6-9 | 369 (35.9%) | 181 (42.0%) | 0 | |
| 10-13 | 288 (28.0%) | 81 (18.8%) | 0.29 (0.09 - 0.51) | 0.003 |
| Missing | 22 (2.1%) | 18 (4.2%) | 0.00 (-0.16 - 0.00) | 0.616 |
| **Type of school qualification, n (%)** | | | | |
| No school leaving qualification | 7 (0.7%) | 9 (2.1%) | 0.00 (0.00 - 0.00) | 0.984 |
| Secondary school qualification (class 8/9) | 49 (4.8%) | 20 (4.6%) | 0.00 (0.00 - 0.00) | 0.996 |
| Secondary school qualification (class 10) | 190 (18.5%) | 104 (24.1%) | -0.10 (-0.31 - 0.00) | 0.187 |
| A-Levels (class 12 or 13) | 776 (75.6%) | 288 (66.8%) | 0 | |
| Missing | 5 (0.5%) | 10 (2.3%) | 0.00 (-0.01 - 0.00) | 0.970 |
| **Monthly net equivalent income, n (%)** | | | | |
| <1,000€ | 79 (7.7%) | 74 (17.2%) | -0.27 (-0.50 - -0.01) | 0.021 |
| 1,000 – 1,999€ | 234 (22.8%) | 126 (29.2%) | -0.08 (-0.28 - 0.00) | 0.243 |
| 2,000 – 2,999€ | 294 (28.6%) | 120 (27.8%) | 0 | |
| 3,000 – 3,999€ | 189 (18.4%) | 49 (11.4%) | 0.16 (0.00 - 0.39) | 0.098 |
| 4,000 – 4,999€ | 143 (13.9%) | 33 (7.7%) | 0.09 (0.00 - 0.33) | 0.238 |
| ≥5,000€ | 44 (4.3%) | 10 (2.3%) | 0.00 (0.00 - 0.09) | 0.897 |
| Missing | 44 (4.3%) | 19 (4.4%) | 0.00 (-0.04 - 0.00) | 0.958 |
| **Sexlife, n (%)** | | | | |
| Content | 717 (69.8%) | 206 (47.8%) | 0 | |
| Discontent | 148 (14.4%) | 108 (25.1%) | -0.31 (-0.52 - -0.07) | 0.006 |
| Sex doesnt matter right now | 12 (1.2%) | 14 (3.2%) | 0.00 (0.00 - 0.00) | 0.990 |
| Missing | 150 (14.6%) | 103 (23.9%) | -0.26 (-0.48 - -0.02) | 0.014 |
| **Number of sex partners within the last 6 months, n (%)** | | | | |
| 0 | 2 (0.2%) | 8 (1.9%) | 0.00 (0.00 - 0.00) | 0.995 |
| 1 | 3 (0.3%) | 21 (4.9%) | -0.26 (-0.50 - 0.00) | 0.047 |
| 2-3 | 123 (12.0%) | 134 (31.1%) | -0.76 (-0.98 - -0.54) | 0.000 |
| 4-5 | 164 (16.0%) | 101 (23.4%) | -0.31 (-0.52 - -0.07) | 0.005 |
| 6-10 | 242 (23.6%) | 83 (19.3%) | 0.00 (0.00 - 0.12) | 0.810 |
| 11-20 | 193 (18.8%) | 35 (8.1%) | 0.26 (0.04 - 0.50) | 0.012 |
| >20 | 286 (27.8%) | 47 (10.9%) | 0 | |
| Missing | 14 (1.4%) | 2 (0.5%) | 0.00 (0.00 - 0.00) | 1.000 |
| **Condom use, n (%)** | | | | |
| 0% | 322 (31.4%) | 62 (14.4%) | 0.49 (0.28 - 0.70) | 0.000 |
| 25% | 339 (33.0%) | 113 (26.2%) | 0 | |
| 50% | 154 (15.0%) | 101 (23.4%) | -0.29 (-0.51 - -0.07) | 0.006 |
| 75% | 106 (10.3%) | 55 (12.8%) | 0.00 (-0.18 - 0.00) | 0.702 |
| >95% | 86 (8.4%) | 97 (22.5%) | -0.59 (-0.83 - -0.35) | 0.000 |
| Missing | 20 (1.9%) | 3 (0.7%) | 0.00 (0.00 - 0.00) | 0.975 |
| **Number of sexual intercourses in the last month, n (%)** | | | | |
| 0 | 69 (6.7%) | 63 (14.6%) | -0.07 (-0.31 - 0.00) | 0.310 |
| 1-4 | 442 (43.0%) | 217 (50.3%) | 0 | |
| 5-8 | 232 (22.6%) | 68 (15.8%) | 0.01 (0.00 - 0.23) | 0.427 |
| 9-12 | 104 (10.1%) | 40 (9.3%) | 0.00 (-0.10 - 0.00) | 0.873 |
| >12 | 168 (16.4%) | 38 (8.8%) | 0.12 (0.00 – 0.36) | 0.152 |
| Missing | 12 (1.2%) | 5 (1.2%) | 0.00 (0.00 - 0.00) | 1.000 |
| **Drugs during sex in the last 6 months, n (%)** | | | | |
| Yes | 201 (19.6%) | 130 (30.2%) | -0.32 (-0.51 - -0.10) | 0.002 |
| No | 808 (78.7%) | 289 (67.1%) | 0 | |

| | PrEP users, n (%) | Non-PrEP users, n (%) | β (95% CI) | *P*-value |
|---|---|---|---|---|
| Missing | 18 (1.8%) | 12 (2.8%) | 0.00 (-0.03 - 0.00) | 0.964 |
| **Injecting drugs during sex, n (%)** | | | | |
| Yes, but not in the last 6 months | 2 (0.2%) | 5 (1.2%) | 0.00 (0.00 - 0.00) | 1.000 |
| Yes, 1-3x in the last 6 months | 10 (1.0%) | 4 (0.9%) | 0.00 (0.00 - 0.00) | 1.000 |
| Yes, >3x in the last 6 months | 17 (1.7%) | 6 (1.4%) | 0.00 (0.00 - 0.00) | 1.000 |
| No | 171 (16.7%) | 114 (26.5%) | -0.09 (-0.30 - 0.00) | 0.192 |
| Missing | 827 (80.5%) | 302 (70.1%) | 0 | |
| **Payment for sex in the last 6 months, n (%)** | | | | |
| Yes | 58 (5.6%) | 37 (8.6%) | 0.00 (-0.19 - 0.00) | 0.667 |
| No | 964 (93.9%) | 381 (88.4%) | 0 | |
| Missing | 5 (0.5%) | 13 (3.0%) | 0.00 (-0.07 - 0.00) | 0.920 |
| **Number of syphilis diagnosis in the last 12 months, n (%)** | | | | |
| 0 | 166 (16.2%) | 52 (12.1%) | 0.00 (0.00 - 0.12) | 0.849 |
| 1 | 107 (10.4%) | 42 (9.7%) | 0.00 (-0.06 - 0.00) | 0.939 |
| ≥2 | 11 (1.1%) | 2 (0.5%) | 0.00 (0.00 - 0.00) | 1.000 |
| Missing | 743 (72.3%) | 335 (77.7%) | 0 | |
| **Number of gonorrhoea diagnosis in the last 12 months, n (%)** | | | | |
| 0 | 173 (16.8%) | 65 (15.1%) | 0.00 (-0.14 - 0.00) | 0.747 |
| 1 | 200 (19.5%) | 56 (13.0%) | 0.03 (0.00 - 0.24) | 0.398 |
| ≥2 | 68 (6.6%) | 10 (2.3%) | 0.00 (0.00 - 0.14) | 0.770 |
| Missing | 586 (57.1%) | 300 (69.6%) | 0 | |
| **Number of chlamydia diagnosis in the last 12 months, n (%)** | | | | |
| 0 | 132 (12.9%) | 40 (9.3%) | 0.00 (0.00 - 0.00) | 0.975 |
| 1 | 221 (21.5%) | 59 (13.7%) | 0.00 (0.00 - 0.08) | 0.896 |
| ≥2 | 59 (5.7%) | 2 (0.5%) | 0.10 (0.00 - 0.28) | 0.173 |
| Missing | 615 (59.9%) | 330 (76.6%) | 0 | |
| **Number of hepatitis C diagnosis in the last 12 months, n (%)** | | | | |
| 0 | 20 (1.9%) | 17 (3.9%) | 0.00 (-0.21 - 0.00) | 0.564 |
| 1 | 5 (0.5%) | 1 (0.2%) | 0.00 (0.00 - 0.00) | 1.000 |
| Missing | 1,002 (97.6%) | 413 (95.8%) | 0 | |
| **Gender of sexual partners, n (%) (multiple responses possible)** | | | | |
| Male | 1,027 (100.0%) | 430 (99.8%) | 0.00 (0.00 - 0.00) | 1.000 |
| Female | 44 (4.3%) | 46 (10.7%) | -0.24 (-0.49 - 0.00) | 0.053 |
| Non-binary | 33 (3.2%) | 13 (3.0%) | 0.00 (0.00 - 0.00) | 0.999 |
| Missing | 0 (0%) | 0 (0%) | - | |
| **STI symptoms in the last 12 months, n (%)** | | | | |
| Yes | 352 (34.3%) | 135 (31.3%) | 0.00 (-0.14 - 0.00) | 0.741 |
| No | 673 (65.5%) | 294 (68.2%) | 0 | |
| Missing | 2 (0.2%) | 2 (0.5%) | 0.00 (0.00 - 0.00) | 1.000 |
| **Positive STI test in your life, n (%)** | | | | |
| Syphilis | 288 (28.0%) | 99 (23.0%) | 0.00 (-0.02 - 0.10) | 0.835 |
| Gonorrhoea | 444 (43.2%) | 131 (30.4%) | 0.10 (0.00 - 0.29) | 0.168 |
| Chlamydia | 420 (40.9%) | 101 (23.4%) | 0.39 (0.19 - 0.60) | 0.000 |
| Hepatitis C | 25 (2.4%) | 18 (4.2%) | 0.00 (-0.13 - 0.00) | 0.754 |
| Never | 322 (31.4%) | 177 (41.1%) | 0.00 (0.00 - 0.09) | 0.865 |
